# Infodemic Management Challenges and Training Needs Among Frontline Health Educators in Lagos State Nigeria

**DOI:** 10.64898/2026.04.09.26350557

**Authors:** Abara Erim, Patrick Lansana, Olayinka Badmus, Motunrayo Florence Olanrewaju

## Abstract

Misinformation circulating through digital platforms and community networks increasingly challenges public health communication, particularly in low-and middle-income countries. Frontline health educators play a critical role in addressing misinformation and promoting accurate health information within primary health care systems; however, empirical evidence on their preparedness to manage infodemics remains limited. This study assessed the training needs and response capacity of primary health care health educators in Lagos State, Nigeria. A convergent mixed-methods design was employed across three districts. Quantitative data were collected from 95 health educators using the 30-item Health Educators’ Infodemic Management Training Needs Assessment Questionnaire (HEIM-TNAQ). Qualitative data were obtained through six focus group discussions involving 56 educators and 25 key informant interviews with supervisors and programme managers. Quantitative data were analysed using descriptive statistics and t-tests, while qualitative data were analysed thematically. Participants demonstrated relatively strong knowledge of health misinformation (mean = 71.5), but only moderate decision-response skills (48.6) and low confidence in addressing misinformation (42.5). Integration of misinformation response into routine practice was also limited (46.3), and no significant differences were observed between respondents with or without prior training. Qualitative findings revealed frequent exposure to vaccine rumours, spiritual explanations for illness, and misinformation circulating through social media and community networks. Strengthening infodemic management within primary health care requires practical training, behavioural communication skills, and institutional mechanisms for systematic rumour monitoring and response.

## Introduction

The rapid expansion of digital communication has transformed how communities access, share, and interpret health information. During public health emergencies, this environment facilitates the spread of misinformation, disinformation, and information overload phenomena collectively described as an *infodemic* ^1,2^. Infodemics amplify public confusion, erode trust in health authorities, and undermine adherence to recommended health behaviours. As seen during COVID-19, the infodemic often became as consequential as the pandemic itself, influencing risk perception, health-seeking behaviour, and acceptance of vaccines and other public health interventions ^3,4^.

Frontline health educators, particularly those working within primary health care systems in low-and middle-income countries (LMICs) play a critical role in mediating how communities interpret and act on health information. Yet, despite their centrality, research on their preparedness to manage misinformation remains limited ^5,6^. Much of the existing literature focuses on clinicians, digital responders, or national-level risk communication specialists, leaving a gap in understanding the community-level workforce that interacts most directly with populations ^1,7^. This gap is especially concerning given evidence that misinformation in LMIC contexts often spreads simultaneously through digital channels (e.g., WhatsApp, Facebook) and traditional community structures such as markets, religious gatherings, and local leaders, requiring nuanced, context-responsive approaches to mitigation ^8,9,10^.

Recognizing the growing threat of infodemics, the World Health Organization (WHO) has developed a global competency framework to guide institutions in building a skilled infodemic management workforce ^11–13^. The framework outlines core domains necessary for effective practice, including surveillance of misinformation trends, community engagement, risk communication, and the design and evaluation of interventions. Complementing this, WHO and partners have launched multiformat infodemic management training programmes, simulation exercises, and digital learning modules to enhance health workers’ competencies worldwide ^13,14^. Despite these global advances, the extent to which such competencies exist and where training gaps persist among frontline community-based health educators in LMIC settings remains poorly understood^15,16^.

Moreover, recent WHO and academic work highlights that effective infodemic management must extend beyond crisis response to become integrated into routine public health practice ^1,14^. This includes proactive social listening, strengthening community trust, communicating uncertainty transparently, and building systems that support rapid rumour detection and response. Understanding the current capabilities, challenges, and training needs of frontline health educators is therefore essential for strengthening national preparedness and community resilience.

Given these gaps, the present study examines the infodemic-related competencies, experiences, and training needs of frontline health educators in Nigeria. By combining quantitative assessments with qualitative insights, this research contributes evidence needed to inform workforce development, curriculum design, and system-level strategies for building a resilient health promotion workforce capable of managing infodemics effectively.

## Materials and Methods

### Study Design

This study employed a mixed-methods design, beginning with a qualitative phase to explore frontline health educators’ experiences with health misinformation, followed by a quantitative phase to measure the prevalence of identified capacity gaps across a wider sample of practitioners. Insights from the qualitative phase informed the development and refinement of the survey instrument used in the quantitative assessment.

### Study Setting and Participants

The study was conducted in Lagos State, Nigeria, across three purposively selected districts representing diverse population characteristics and primary health care (PHC) service structures. These districts manage routine health promotion, community engagement, and risk communication activities through PHC facilities and LGA-level health education units.

The target population consisted of frontline health educators and supervisors responsible for delivering community engagement and health promotion activities. A total of 120 participants were included in the study. The quantitative component involved 95 health educators, while the qualitative component comprised six focus group discussions (FGDs) with 56 health educators and 25 key informant interviews (KIIs) with supervisors and programme managers overseeing PHC-level communication and community engagement activities. Some participants participated in both the qualitative and quantitative assessment.

Participants represented multiple cadres, including health educators, community health officers, surveillance officers, programme managers, and LGA-level health education coordinators. The flow of participants across the quantitative and qualitative components of the study is illustrated in **Figure 1**.

**Figure 1.**
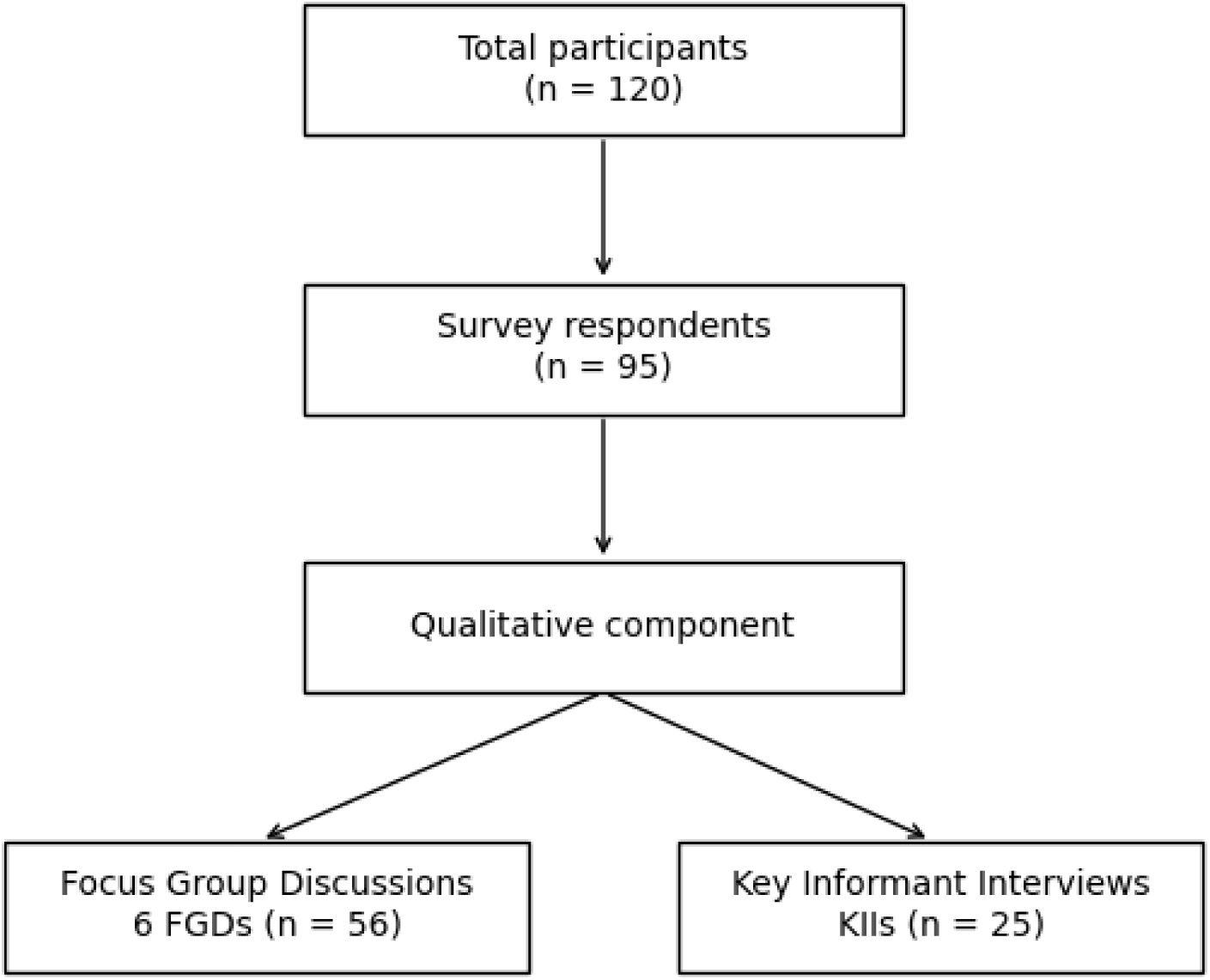
Participant flow diagram for the mixed-methods study. The study included 120 participants. The quantitative component consisted of 95 health educators who completed the training needs assessment survey. The qualitative component comprised six focus group discussions (FGDs) with 56 health educators and 25 key informant interviews (KIIs) with supervisors and programme managers.

### Sampling Procedures Qualitative Phase

A purposive sampling strategy was used to recruit FGD and KII participants from three districts. This approach ensured representation across professional roles, years of experience, and districts. Survey respondents who indicated willingness to participate in further interviews were prioritized for inclusion.

Each FGD consisted of 8–12 participants, while KIIs were conducted with supervisors and district-level decision-makers responsible for oversight of health education services.

### Quantitative Phase

A convenience sampling approach was used to recruit health educators for the quantitative assessment. Eligible participants included individuals actively engaged in community outreach, health education, or health promotion activities within PHC facilities across the districts. All available personnel meeting inclusion criteria during scheduled data collection periods were invited to participate.

### Data Collection Procedures Qualitative Component

FGDs and KIIs were conducted using semi-structured interview guides developed from preliminary consultations with district health officials and existing literature on misinformation. Discussions explored common misinformation themes, sources and channels of spread, response practices, challenges, and perceived training needs.

All sessions were conducted in English, audio-recorded with consent, transcribed verbatim, and supplemented with field notes documenting contextual observations.

### Quantitative Component

The quantitative phase used the Health Educators’ Infodemic Management Training Needs Assessment Questionnaire (HEIM-TNAQ), a 30-item survey developed based on qualitative insights.

The instrument assessed five domains:

1. Knowledge of health misinformation
2. Decision-response skills
3. Attitudes toward addressing misinformation
4. Confidence in responding to misinformation
5. Integration of misinformation activities into routine practice

Knowledge and decision-response items were scored dichotomously (1 = correct, 0 = incorrect), while Likert scales (1–5) were used for attitudes, confidence, and integration, with reverse-coded items adjusted during analysis. Surveys were self-administered during facility meetings and staff gatherings across the three districts.

## Data Analysis

### Qualitative Analysis

Transcripts from FGDs and KIIs were analysed using thematic analysis following iterative coding, theme development, and cross-case comparison. Themes reflected the types and drivers of misinformation, response practices, perceived competencies, challenges, and training and system-level needs. Qualitative findings informed refinement of survey items and supported triangulation with quantitative results.

### Quantitative Analysis

Survey data were cleaned and analysed to produce descriptive statistics (means, standard deviations, frequency distributions) for each domain. Mean domain scores were computed according to the survey scoring guide.

Independent samples t-tests compared domain scores between participants with prior training in health communication, misinformation, behavioural intelligence, or infodemic management and those without such training. Internal consistency for each domain and the overall instrument was assessed using Cronbach’s alpha.

### Mixed-Methods Integration

Integration occurred at the interpretation stage, where quantitative patterns were contextualized using qualitative insights, allowing identification of convergent, divergent, and complementary findings.

### Ethical Considerations

Ethical approval for the study was obtained from the Babcock University Health Research Ethics Committee (BUHREC) with approval Number BHUREC0126/26/1645. Participant recruitment and data collection were conducted between February, 2026 and April, 2026 across the selected districts in Lagos State.

All participants provided written informed consent prior to participation. Participation was voluntary, and participants were informed of their right to withdraw at any time without consequence. Confidentiality was ensured through anonymization of data and secure data handling procedures.

The study did not involve minors; therefore, parental or guardian consent was not required.

## Results

### Participant Demographics

A total of 120 participants took part in the study. Of these, 95 completed the quantitative survey, while 56 participated in focus group discussions and 25 in key informant interviews (some individuals participated in both phases).

The socio-demographic and professional characteristics of the survey respondents are presented in Table 1. The majority of participants were female (64.2%), and most were between 20 and 39 years of age (75.8%). Nearly half of the respondents were health educators (48.4%), followed by health promotion officers (31.6%). Approximately 50% of participants reported having received prior training related to health communication or misinformation, while the remaining respondents had not received such training.

**Table 1.** Socio-Demographic and Professional Characteristics of Participants (n = 95)

| **Characteristic** | **Category** | **n** | **%** |
| --- | --- | --- | --- |
| **Sex** | Female | 61 | 64.2 |
|  | Male | 34 | 35.8 |
| **Age Group (years)** | 20–29 | 36 | 37.9 |
|  | 30–39 | 36 | 37.9 |
|  | 40–49 | 16 | 16.8 |
|  | ≥50 | 7 | 7.4 |
| **Highest Educational Qualification** | Diploma | 13 | 13.7 |
|  | Bachelor's degree | 49 | 51.6 |
|  | Master's degree | 22 | 23.2 |
|  | Doctorate | 11 | 11.6 |
| **Professional Role** | Health Educator | 46 | 48.4 |
|  | Health Promotion Officer | 30 | 31.6 |
|  | Community Health Officer | 14 | 14.7 |
|  | Surveillance Officer | 5 | 5.3 |
| **Years of Experience** | <2 years | 9 | 9.5 |
|  | 2–5 years | 43 | 45.3 |
|  | 6–10 years | 33 | 34.7 |
|  | >10 years | 10 | 10.5 |
| **Previous Training on Health Communication/Misinformation** | Yes | 48 | 50.5 |
|  | No | 47 | 49.5 |
| **Frequency of Community Engagement** | Weekly | 47 | 49.5 |
|  | Bi-weekly | 30 | 31.6 |
|  | Monthly | 12 | 12.6 |
|  | Less than monthly | 6 | 6.3 |

### Infodemic Management Competency Domain

Across the five assessed domains, participants demonstrated varying levels of readiness for infodemic management (Table 2). Overall, knowledge of health misinformation showed the strongest performance, with a mean score of 71.5 (SD = 10.4), indicating generally good conceptual understanding of misinformation, its drivers, and its implications for community health behaviour. However, the wide score range (46–89) suggests that while many respondents possess strong knowledge, a notable subset still lacks foundational understanding.

**Table 2.**
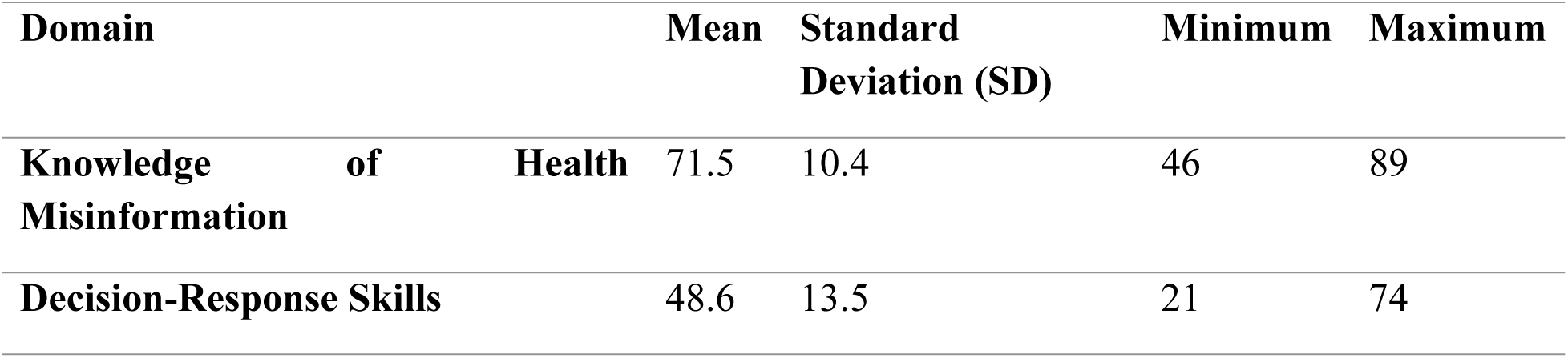
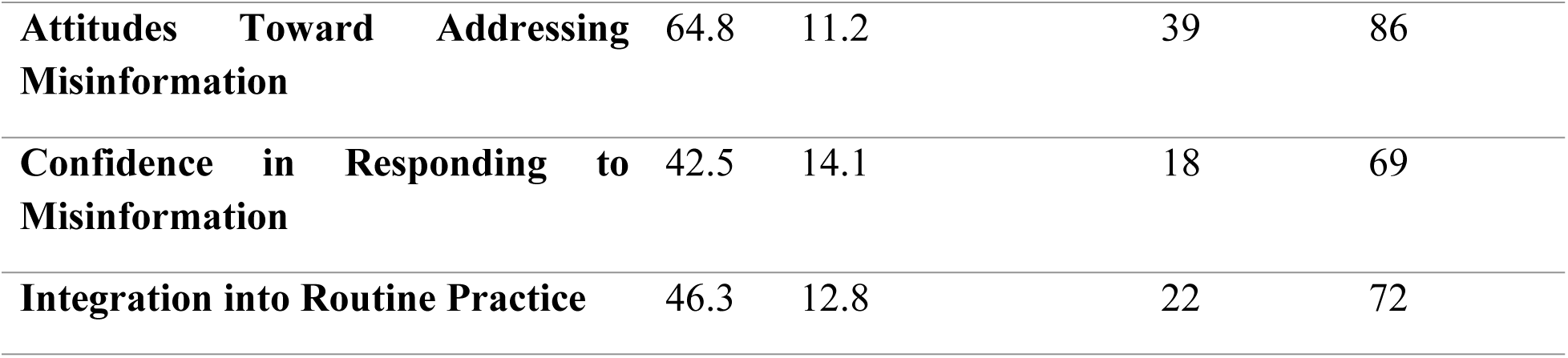
Domain Scores for Infodemic Management Training Needs Assessment (n = 95)

Performance in the decision-response skills domain was moderate, with a mean score of 48.6 (SD = 13.5) and a broad distribution of scores (21–74). This reflects variability in the ability to apply knowledge in practical, real-world scenarios, particularly when engaging with resistant community members or addressing misinformation deeply rooted in cultural beliefs.

Participants expressed generally positive attitudes toward addressing misinformation, as reflected in a mean score of 64.8 (SD = 11.2). Most respondents agreed that addressing misinformation is a key component of their role and recognized the importance of proactive community engagement.

In contrast, confidence in responding to misinformation was comparatively low, with a mean score of 42.5 (SD = 14.1). This suggests that although participants may understand misinformation conceptually and hold positive attitudes toward addressing it, many do not feel adequately equipped or self-assured when required to respond in real-time community contexts.

The integration of misinformation response into routine practice also scored modestly, with a mean of 46.3 (SD = 12.8). This indicates limited systematic incorporation of misinformation monitoring, documentation, and reporting into daily responsibilities likely reflecting broader structural or institutional constraints rather than individual unwillingness.

### Comparison of Domain Scores by Training Exposure

Contrary to expectations, respondents who reported previous training in health communication or misinformation-related content did not perform significantly better than those without such training across any of the assessed domains (Table 3). In all five domains, p-values exceeded 0.05, indicating no statistically significant differences between the two groups.

**Table 3.** Comparison of Domain Scores by Previous Training Exposure (n = 95)

| **Domain** | **Previous Training Mean (SD)** | **No Previous Training Mean (SD)** | **p-value** |
| --- | --- | --- | --- |
| **Knowledge** | 69.44 (19.55) | 71.63 (16.99) | 0.562 |
| **Decision-Response Skills** | 47.92 (20.23) | 51.77 (19.73) | 0.349 |
| **Attitudes** | 63.54 (7.70) | 64.18 (8.55) | 0.701 |
| **Confidence** | 57.15 (8.08) | 58.37 (7.95) | 0.462 |
| **Integration into Routine Practice** | 57.08 (8.38) | 56.52 (8.60) | 0.749 |

While minor variations were observed, such as slightly higher confidence and integration scores among the trained group, these differences were not large enough to reach statistical significance. This pattern suggests that prior training may have been either insufficiently comprehensive, outdated, lacking practical components, or not specifically targeted toward infodemic management competencies. It may also reflect the need for more tailored, skill-focused capacity-building interventions that address the realities of misinformation within community settings.

### Reliability Analysis

The reliability analysis of the HEIM-TNAQ instrument demonstrated strong internal consistency (Table 4). Cronbach’s alpha values for the five individual domains ranged from 0.76 to 0.84, indicating acceptable to high reliability across domains. The overall scale achieved a Cronbach’s alpha of 0.88, reflecting excellent internal consistency.

**Table 4.** Reliability Analysis of HEIM-TNAQ Domains.

| Domain | Number of Items | Cronbach's Alpha ($\alpha$) |
| --- | --- | --- |
| Knowledge of Health Misinformation | 6 | 0.78 |
| Decision-Response Skills | 6 | 0.82 |
| Attitudes Toward Addressing Misinformation | 6 | 0.76 |
| Confidence in Responding to Misinformation | 6 | 0.84 |
| Integration into Routine Practice | 6 | 0.80 |
| Overall Scale | 30 | **0.88** |

**Table 5.**
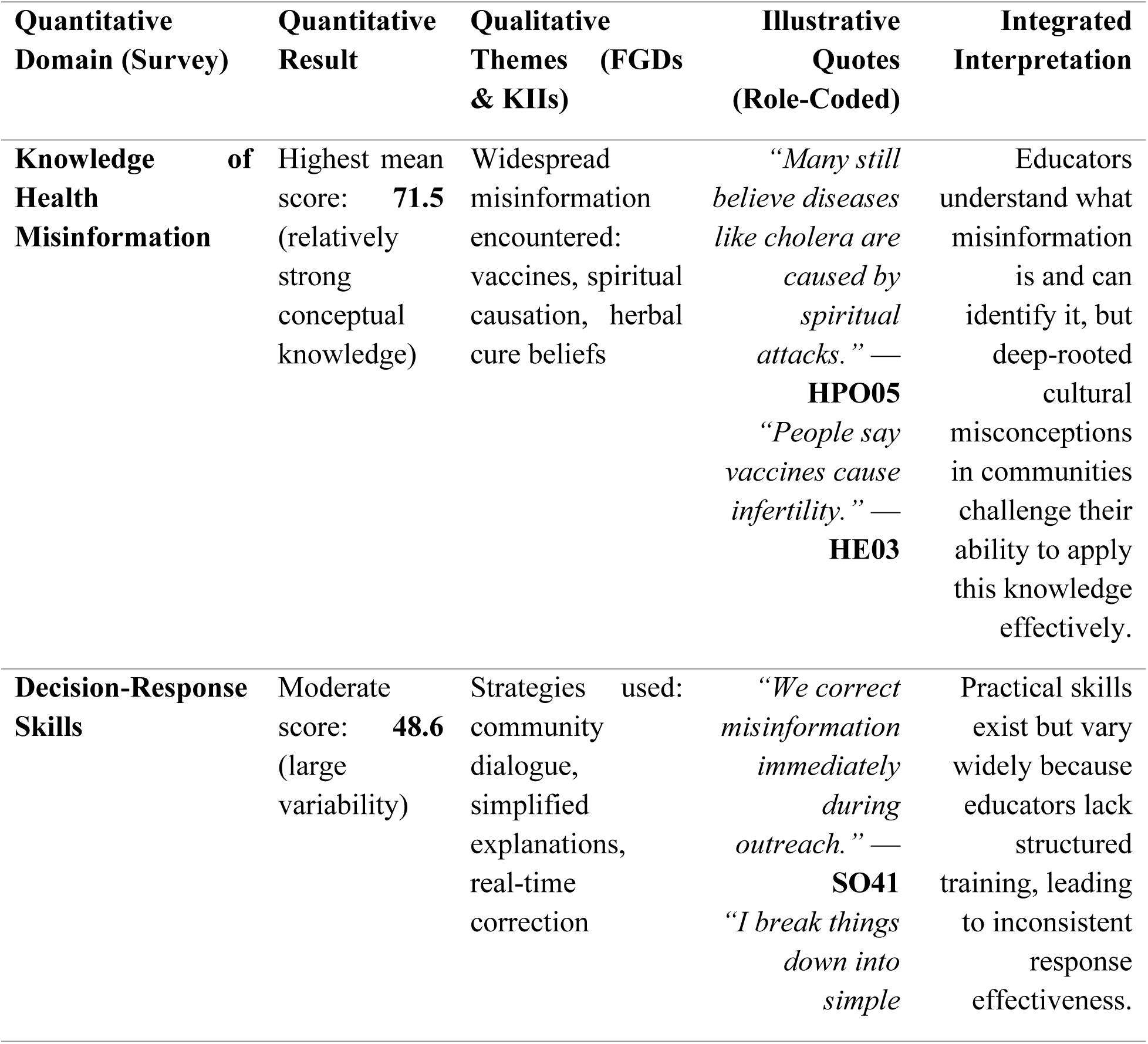
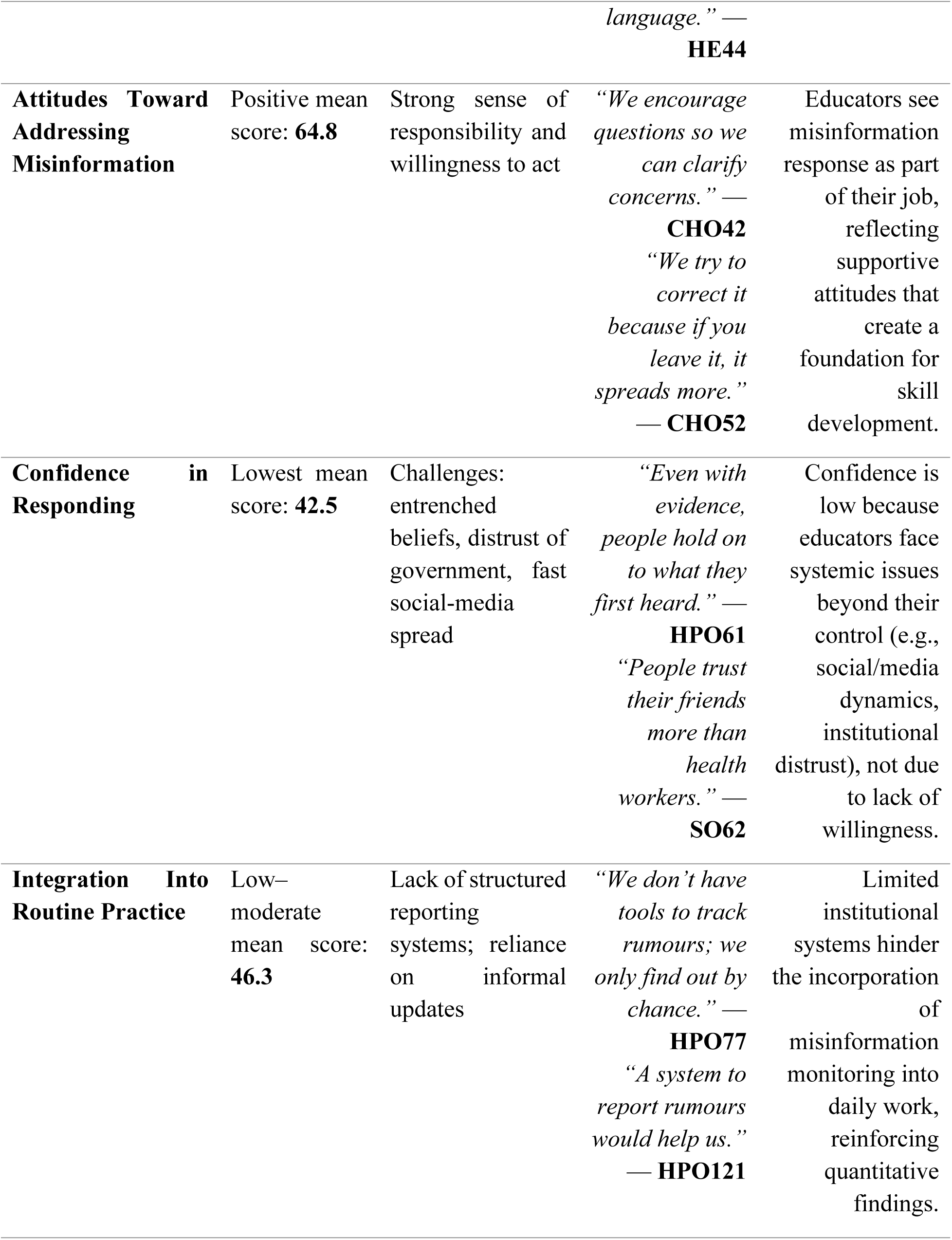
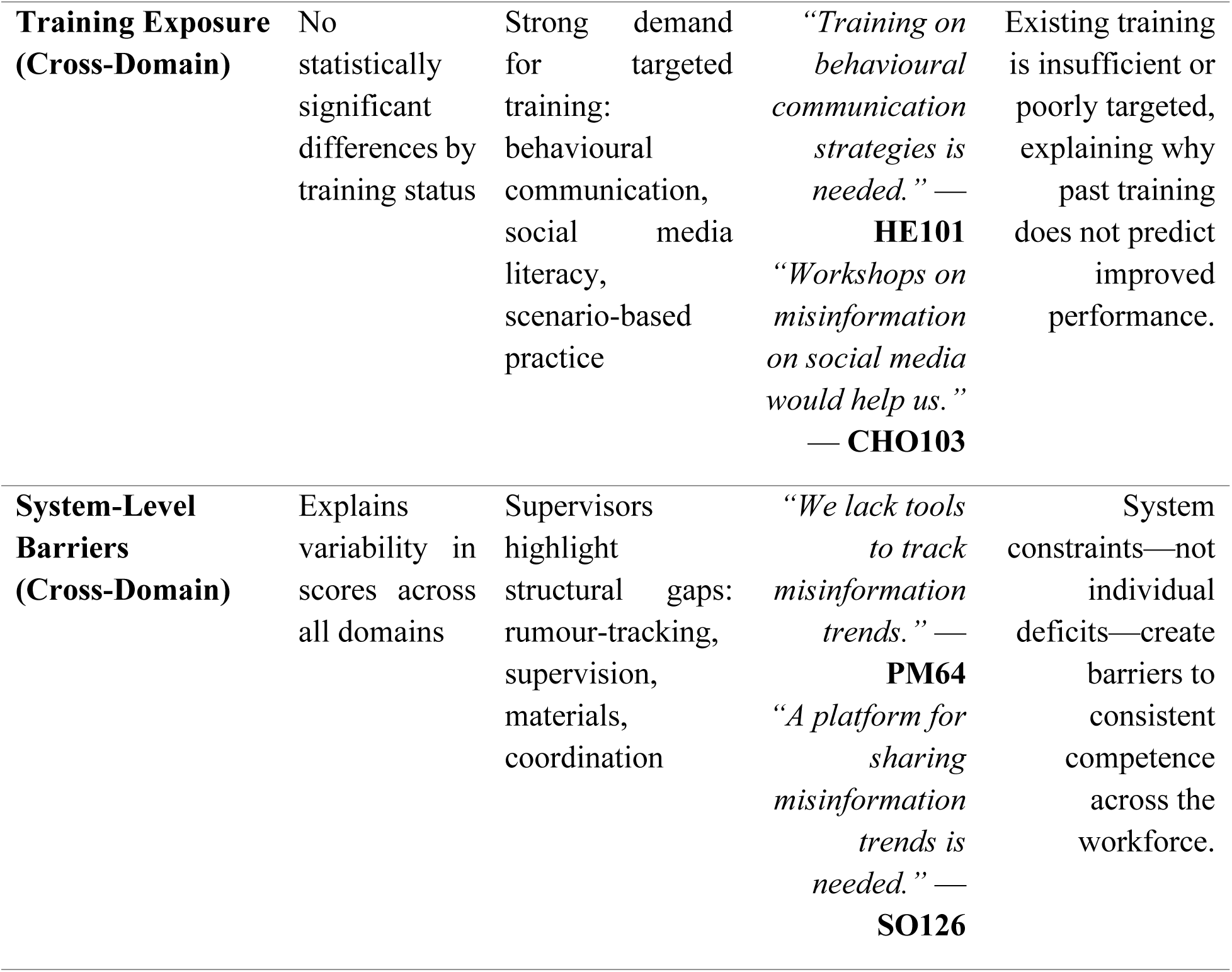
Joint Display of Quantitative and Qualitative Findings. This joint display illustrates how qualitative insights help explain quantitative patterns observed across the five assessed domains of infodemic management capacity.

These results suggest that the instrument reliably captures the multidimensional constructs associated with infodemic management capacity, including knowledge, practical skills, attitudes, confidence, and routine integration. The strong psychometric performance supports its suitability for future assessments and capacity-building evaluations within similar health education contexts.

### Qualitative Findings

The qualitative component of the study explored the lived experiences of frontline health educators and supervisors across three districts in Lagos State regarding the types of misinformation encountered, sources of rumours, response strategies, challenges, and training or system-level needs. Five major themes emerged from the analysis.

### Theme 1: Types of Health Misinformation Circulating in Communities

Participants reported encountering a diverse range of misinformation during routine community engagement. These misconceptions reflected both long-standing cultural narratives and more recent digitally mediated rumours, creating a complex infodemic landscape that frontline workers must continually navigate.

A central theme was vaccine-related misinformation, particularly beliefs linking vaccination to infertility, population control, and harmful side effects. These rumours were widespread and emotionally impactful, especially among caregivers. A Health Educator highlighted the deeply entrenched nature of infertility myths, stating:

*“A very common misconception is that vaccines can make young women infertile. Mothers tell me they heard it from a neighbour or a relative.”* — **HE03**

Another health worker emphasized the persistence of this narrative:

*“The rumour about vaccines causing infertility is deeply rooted. Mothers repeat it confidently, like it’s already confirmed.”* — **HPO09**

Related misinformation positioned vaccines as tools of external control or manipulation, often framed within political or geopolitical distrust:

*“People believe vaccines are part of a plan to reduce the population. They say outsiders want to reduce the number of Africans.”* — **HE04**

Beyond vaccines, educators frequently encountered misconceptions about disease causation, often rooted in spiritual or supernatural explanations. These narratives were particularly common for illnesses such as cholera and malaria. One Health Promotion Officer explained:

*“Many still believe diseases like cholera are not caused by dirty water but by spiritual attacks. They think someone has cast a spell.”* — **HPO05**

A Community Health Officer described how spiritual interpretations delay appropriate care:

*“People go to prayer houses first because they think it’s a spiritual attack. By the time they come to the clinic, the situation is worse.”* — **CHO06**

Another prominent misinformation category involved overreliance on herbal remedies. Participants noted that many community members view hospital treatment with suspicion and prefer local mixtures or herbs:

*“In almost every community, people insist herbs cure everything. They say hospitals give ‘strong drugs’ that cause problems.”* — **SO08**

Frontline workers also consistently mentioned outbreak-related misinformation, particularly denial of disease outbreaks or claims that epidemics were fabricated:

*“When an outbreak happens, the first rumour is that it’s fake. People say, ‘We’ve never seen anyone with it,’ or that the government created it to get funds.”* — **HE02**

Such narratives often stem from broader distrust in formal health systems:

*“People insist the disease isn’t real. They say it’s something the government exaggerated to scare people.”* — **HPO01**

Overall, the misinformation landscape described by the participants reveals a blend of cultural belief systems, distrust in institutions, and digitally amplified rumours. This combination creates layered challenges for frontline educators who must respond not only to medical misunderstandings but also to deeply embedded social narratives and rapidly circulating digital misinformation.

### Theme 2: Sources and Channels of Misinformation

Participants across all three districts identified a diverse set of pathways through which misinformation spreads within their communities. These channels included a blend of digital platforms, informal social spaces, and community authority structures, illustrating the multiple layers of the local information environment. The interplay between these sources often enabled misinformation to circulate rapidly and gain credibility before health workers could intervene.

### Digital Platforms as Major Accelerators

The most frequently cited source of misinformation was WhatsApp, particularly community and family group chats where messages circulate unchecked. Frontline workers consistently described WhatsApp as a “primary driver” of rumours due to its rapid forwarding culture and the perceived authenticity of audio or video messages.

*“WhatsApp groups are probably the biggest culprit now. Messages come in with voice notes and everyone forwards them without checking.”* — **CHO30**

Similarly, Facebook was identified as a major channel through which misleading posts, images, or pseudo-medical advice gain traction:

*“Most rumours spread through Facebook posts. People see something with a lot of comments and assume it’s true.”* — **CHO32**

Digital misinformation often blended with existing community fears, making it especially influential.

### Informal Social Spaces as Rumour Hotspots

Participants also described markets as vibrant environments for rumour circulation. Because markets are daily gathering points, unverified information spreads quickly through casual conversations:

*“A lot of rumours start in the markets. People talk while buying food and before you know it, the entire market is repeating the same thing.”* — **HE21**

Community meetings were another common offline channel. In these settings, misinformation often becomes entrenched when repeated by individuals considered credible or influential:

*“In community meetings, if an influential person says something, many people believe it immediately.”* — **CHO22**

This highlights how social hierarchies amplify the spread of false information.

### Traditional and Religious Structures as Credibility Amplifiers

Many health educators emphasized that religious gatherings and statements made by faith leaders significantly influenced public beliefs. Because religious figures hold substantial authority, their words are often accepted without question:

*“When a religious leader mentions something casually, people treat it like verified truth.”* — **HPO23**

Similarly, traditional leaders and elders were frequently cited as unintentional vectors of misinformation:

*“Most rumours spread through traditional leaders. Once they repeat it, people don’t question it.”* — **CHO33**

These structures act as both stabilizers of community norms and accelerators of misinformation, depending on the accuracy of the information being shared.

### Interplay Between Digital and Community Channels

Participants repeatedly described how misinformation often originates online, spreads rapidly through WhatsApp or Facebook, and then becomes amplified in physical community spaces such as markets, churches, mosques, and meetings. This creates a reinforcing cycle where rumours feel legitimate because they are encountered in multiple contexts.

One surveillance officer captured this dynamic:

*“Most rumours spread through WhatsApp messages, but then people confirm them with neighbours or leaders, so it becomes even stronger.”* — **SO34**

This blending of digital and interpersonal spread pathways makes rumours harder to challenge, as community members rely on both their social networks and online ecosystems for information validation.

### Theme 3: Current Strategies for Responding to Health Misinformation

Frontline health educators described employing a range of interpersonal and community-based strategies to address misinformation encountered during outreach activities. These strategies demonstrate both adaptability and a strong commitment to community engagement, despite limited formal training or institutional tools for addressing misinformation.

### Creating Dialogue and Encouraging Community Questions

A central approach described across FGDs and KIIs was the deliberate creation of open dialogue spaces where community members could express doubts, ask questions, and discuss misinformation without fear of judgment. Participants emphasized that opening the floor for discussion often reveals the underlying concerns driving rumours.

*“We encourage people to ask questions during community dialogue. Sometimes misinformation comes from misunderstanding.”* — **CHO42**

*“During our health talks, we let them speak freely first, then we clarify their concerns one by one.”* — **HE47**

This strategy reflects an understanding that misinformation often thrives in information gaps, making dialogue a powerful corrective tool.

### Simplifying Complex Health Information

Participants frequently noted the importance of simplifying biomedical information to make explanations accessible, relatable, and culturally grounded. This was especially crucial when addressing scientific or clinical concepts such as vaccine safety, disease transmission, and treatment guidelines.

*“I try to break things down into simple language. If you use too much medical terminology, people get confused.”* — **HE44**

*“We give examples from everyday life so they can understand why the information they heard isn’t correct.”* — **SO48**

Simplification helps bridge literacy gaps and reduces the distance between formal health systems and community knowledge systems.

### Real-Time Correction of Misinformation During Outreach

Many educators reported correcting misinformation on the spot during community sessions whenever harmful or widely circulated rumours surfaced.

*“When we hear misinformation, the first thing we do is correct it immediately, especially during health talks.”* — **SO41**

*“We try to correct the information there and then because if you leave it, it spreads more.”* — **CHO52**

This proactive approach reflects the urgency educators feel when confronting misinformation that can quickly escalate or influence harmful behaviours.

### Escalation and Reporting of Serious Rumours

Participants also described escalating dangerous or fast-spreading misinformation to their supervisors or LGA-level communication teams. Although formal reporting systems were limited, many educators relied on informal channels to ensure rumours were addressed as part of broader communication efforts.

*“If the rumour is serious or spreading too fast, I report it to my supervisor so it can be escalated.”* — **SO46**

*“We sometimes report serious rumours to our supervisors because they may need to address it at a higher level.”* — **HPO50**

This practice highlights the perceived importance of linking frontline insights with broader health system responses.

### Using Community-Based Examples and Testimonies

Some educators described using real stories, local examples, or testimonies from trusted community figures to correct misinformation. Although not always formalized, these approaches were believed to resonate strongly with residents.

*“Sometimes I call someone the community trusts to help explain. When it comes from them, people listen more.”* — **HE49**

This demonstrates how educators strategically embed communication within social norms and trusted networks.

### Theme 4: Challenges in Addressing Misinformation

Frontline health educators across the three districts described significant challenges that hinder their ability to effectively counter misinformation during community engagement. These challenges were deeply interconnected, involving community-level belief systems, structural limitations, and rapid information dynamics shaped by digital media.

### Deeply Entrenched Beliefs and Cultural Narratives

One of the most frequently mentioned barriers was the difficulty of overturning long-standing cultural or spiritual beliefs. Educators explained that once a rumour resonates with pre-existing cultural explanations, it becomes extremely resistant to correction.

*“Once a rumour spreads widely, it becomes very difficult to change people’s minds. Even when you show them evidence, they hold on to what they first heard.”* — **HPO61**

This challenge was particularly acute for diseases perceived as spiritual in origin, where biomedical explanations were often dismissed:

*“Some people think the government cannot be trusted, so they automatically reject any information coming from official sources.”* — **HPO64**

### Low Trust in Government and Formal Health Systems

Participants consistently emphasized low institutional trust as a key factor shaping how communities interpret information. Many residents perceive government messaging with suspicion, leading them to preferentially trust informal sources such as friends, relatives, or religious figures.

*“People trust their friends more than they trust health workers. Even when we explain the facts, they still go back to what their neighbour told them.”* — **SO62**

*“Some community members distrust government information, so they don’t believe us when we bring messages from the health authorities.”* — **CHO66**

This distrust creates an uphill battle for frontline educators, whose authority is often undermined by broader societal skepticism.

### Rapid Spread of Misinformation Through Social Media

Participants described the speed of misinformation spread, especially through WhatsApp and Facebook, as overwhelming. By the time educators reach the community, rumours have often circulated widely, making correction more difficult.

*“Social media spreads misinformation faster than we can correct it. By the time we reach the community, the rumour has already gone far.”* — **HPO68**

This speed mismatch leaves educators constantly reacting, rather than proactively addressing emerging rumours.

### Absence of Tools and Systems for Monitoring Rumours

Educators across roles noted the lack of structured systems for rumour tracking or timely updates on circulating misinformation. Many only discover new rumours during field visits, limiting their ability to prepare responses.

*“We don’t have tools to track the kinds of rumours circulating. We only find out by chance.”* — **HPO77**

Without a mechanism for systematic rumour detection, misinformation often spreads unchecked until it becomes deeply embedded.

### Social Influence of Trusted Figures

Participants stressed the powerful role of community influencers, including religious and traditional leaders. When these figures repeat misinformation, it becomes extremely challenging for health workers to counter it:

*“If a religious leader says something, people believe it more than what we as health workers say.”* — **PM70**

This dynamic reinforces misinformation and makes corrective communication more sensitive and complex.

### Emotional Resistance and Defensive Reactions

Some educators noted that confronting misinformation directly sometimes leads to defensive or emotional reactions from community members, particularly when beliefs are tied to identity, spirituality, or distrust.

*“Some people get defensive when you try to correct them. They feel you’re attacking their belief or calling them ignorant.”* — **HE79**

This creates an additional communication barrier that requires sensitivity and advanced communication skills.

### Theme 5: Training and Support Needs

Participants across all three districts emphasized significant gaps in both formal training and institutional support systems, expressing a strong need for capacity-building interventions tailored to the realities of misinformation in community settings. Their responses highlighted the desire for practical, structured, and continuous training, as well as organisational mechanisms that would strengthen their effectiveness when responding to misinformation.

### Need for Behavioural Communication and Interpersonal Skills Training

Many respondents noted that existing training focused primarily on general health education rather than the specific behavioural competencies needed to address misinformation. They expressed the need for training that equips them to manage resistance, engage sceptical audiences, and respond to emotionally charged encounters.

*“Training on behavioural communication strategies would help us a lot because many people hold on to their beliefs strongly.”* — **HE101**

*“We need to learn how to engage resistant community members without making them defensive.”* — **SO105**

Such training is viewed as essential for navigating complex interpersonal and sociocultural dynamics that influence belief systems at the community level.

### Demand for Social Media and Digital Literacy Skills

Participants frequently highlighted the lack of training on how misinformation spreads in digital environments. Given that WhatsApp and Facebook were identified as major drivers of rumours, many felt unprepared to respond effectively to misinformation originating online.

*“Workshops on how misinformation spreads on social media would really help us. Most rumours now start online.”* — **CHO103**

*“We need training on how to use digital platforms to counter misinformation because we are not taught that.”* — **HE113**

This reflects a clear training gap between the digital nature of emerging misinformation and the primarily analog tools available to frontline educators.

### Need for Practical, Scenario-Based Learning

Participants repeatedly stressed that training should be applied, hands-on, and context-specific. Many indicated that while theoretical knowledge is helpful, practical simulations would better prepare them to handle real misinformation encounters.

*“Practical sessions where we practice responding to misinformation scenarios would help us a lot.”* — **CHO114**

*“We need training where we can role-play situations because that’s when you learn what to say.”* — **SO119**

This desire for active learning reflects educators’ recognition that misinformation response requires both skill and confidence—qualities strengthened through rehearsal and practice.

### Call for Regular Refresher Training

Across cadres, participants emphasized the importance of regular and continuous training opportunities, rather than one-off workshops. They stressed that misinformation evolves rapidly and that capacity-building should keep pace with emerging rumours and community dynamics.

*“Regular refresher training on communication skills is important. Everything changes so fast.”* — **HPO109**

*“We need ongoing training because new misinformation keeps coming up.”* — **HE102**

Regular training is seen as necessary not only for knowledge updates but also for maintaining confidence and preparedness among health educators.

### Need for Structured Rumour-Reporting and Monitoring Systems

Participants described the absence of institutional systems that would support more systematic responses to misinformation. Many educators currently rely on informal channels when reporting dangerous rumours.

*“A system to report rumours from communities would help us manage misinformation better.”* — **HPO121**

*“We need a platform for sharing misinformation trends so that everyone is aware of what is circulating.”* — **SO126**

A formal rumour-tracking mechanism—embedded in the PHC system—was identified as a critical support need, enabling better coordination and timely response across stakeholders.

### Desire for Peer-Learning and Supervisory Support

Frontline workers also noted the need for peer-learning sessions, where health educators can share experiences, discuss challenges, and learn from one another. In addition, they emphasized the importance of supervisory presence and support during difficult engagements.

*“Regular peer learning meetings among health educators would really help us share strategies.”* — **HE127**

*“Support from supervisors during difficult community engagements makes a big difference.”* — CHO133

These insights highlight that misinformation response is not only an individual competency but also a team-supported and system-supported function.

### Integration of Quantitative and Qualitative Findings

The integration of quantitative and qualitative findings (Tabe 5) provides a comprehensive understanding of the infodemic management capacities of frontline health educators and supervisors across the three districts in Lagos State. While the quantitative results highlight strengths and gaps across the five assessed domains, the qualitative findings help explain *why* these patterns emerged.

The high knowledge scores align with educators’ extensive exposure to misinformation in their daily work. Qualitative narratives showed they routinely encounter vaccine myths, spiritual explanations for illness, and misinformation about herbal remedies. These experiences reinforce their conceptual understanding, explaining the strong performance in the knowledge domain.

Despite strong knowledge and attitudes, practical decision-response skills showed only moderate levels. Qualitative insights illustrate that while educators use strategies such as dialogue, simplified explanations, and real-time corrections, these responses vary in effectiveness depending on the context. Entrenched beliefs, emotional resistance, and the influence of trusted community figures reduce the impact of these efforts, helping explain the variability in skills scores.

Confidence was the lowest-scoring domain and the qualitative findings clarify why. Educators described feeling overwhelmed by the rapid spread of misinformation on social media, frustrated by persistent distrust in government institutions, and challenged by deeply held cultural beliefs. These contextual barriers diminish educators’ confidence even when they know the correct information and want to respond effectively.

Similarly, low integration scores reflect systemic challenges rather than personal shortcomings. The absence of structured rumour-tracking mechanisms, limited access to communication materials, and lack of coordinated supervisory support were frequently mentioned in interviews. These structural gaps restrict the incorporation of misinformation monitoring and response into routine practice, mirroring the quantitative findings.

Finally, the absence of statistically significant differences between trained and untrained participants is explained by the qualitative data, which showed that existing trainings are infrequent, generic, and not tailored to infodemic management. Participants consistently called for practical, scenario-based, and digital-literacy-focused training, suggesting that current programmes do not adequately prepare educators for real-world misinformation challenges.

Overall, the integrated findings reveal that while knowledge and motivation are strong among health educators, structural constraints, entrenched cultural beliefs, and inadequate training limit their ability to apply skills confidently and consistently. Strengthening infodemic management capacity will therefore require not only individual-level training but also system-level investments in tools, supervision, and coordinated communication processes.

## Discussion

This mixed-methods study examined the infodemic management capacities of frontline health educators and supervisors across three districts in Lagos State, Nigeria. The integrated findings reveal a workforce that demonstrates strong knowledge and positive attitudes, but limited practical skills, low confidence, and inadequate structural support for addressing misinformation. These findings align with and extend existing literature on the challenges faced by health workers in managing evolving information environments.

Quantitatively, participants showed high knowledge of health misinformation concepts, which is consistent with global evidence indicating that health workers often possess foundational awareness of misinformation trends due to their frontline experience during health crises. A systematic review by Abuhaloob et al. (2024) similarly found that frontline personnel across multiple countries were highly aware of misinformation circulating during outbreaks, largely because they directly confronted the consequences in community settings ^17^. Qualitative findings in the present study reinforce this observation, showing that educators routinely encounter entrenched narratives such as vaccine infertility myths and spiritual explanations for disease which deepen their familiarity with misinformation patterns. However, the persistence of such beliefs in Nigerian communities mirrors global trends in which cultural, social, and religious explanations often compete with biomedical narratives, as highlighted in the Lancet Public Health framework on managing information ecosystems ^1^. This suggests that knowledge alone is insufficient to counter deeply rooted misinformation.

Despite strong knowledge and positive attitudes, participants demonstrated moderate decision-response skills, with qualitative data explaining this as a consequence of relying on spontaneous, unstructured communication strategies during community engagements. This aligns with findings from learning intervention reviews indicating that health workers often lack systematic, scenario-based training required to respond effectively to misinformation under pressure ^18^. The difficulties educators described, such as simplifying complex information, responding in real time, and adjusting explanations to audience needs are widely documented challenges in infodemic contexts. For example, Ishizumi et al. (2024) highlight that misinformation thrives partly because health workers are not equipped with communication techniques that integrate behavioral insights or adapt to rapidly evolving digital information flows^1^.

One of the most striking quantitative findings was the low confidence score, a trend that is extensively supported by international evidence. Frontline workers globally have reported feeling overwhelmed during infodemics due to the speed at which misinformation spreads, high emotional stakes during community interactions, and the erosion of public trust in health systems. A WHO storytelling report on the COVID-19 infodemic found that frontline workers across 41 countries frequently felt ill-prepared and emotionally strained when countering misinformation, largely due to community skepticism and the overwhelming volume of conflicting information online^13^. The present study’s qualitative findings echo this, showing that distrust in government messages, deference to religious or traditional leaders, and rapid social-media-driven rumor amplification significantly undermine educators’ confidence. These insights further support Abuhaloob et al.’s (2024) conclusion that infodemic pressures can diminish health workers’ sense of efficacy, especially in settings where digital misinformation spreads faster than corrective communication can keep pace ^17^.

Additionally, integration of misinformation response into routine practice was low, and qualitative data attributed this to systemic weaknesses. Participants consistently described the absence of rumor-tracking tools, limited supervisory support, and lack of coordinated communication structures. These constraints echo findings from the systematic review by Abuhaloob et al. (2024), which revealed that many countries lacked institutionalized infodemic management systems, relying instead on ad hoc or temporary approaches during crises ^17^. The Lancet Public Health framework similarly stresses the need to embed infodemic management into routine public health functions, rather than restricting it to emergency responses ^1^ The results from Lagos State therefore reinforce calls for structural reforms, including digital listening systems, clear reporting procedures, and integrated communication workflows to strengthen health system resilience to misinformation.

Notably, the study found no significant differences in domain scores between trained and untrained participants, suggesting that existing training initiatives are insufficient. This mirrors global findings showing that many learning interventions in health emergencies are largely theoretical, lack practical components, and are often not tailored to misinformation dynamics ^18,19^. The WHO’s global infodemic storytelling analysis also highlights that traditional health communication training does not adequately prepare workers for the emotional, social, and digital complexities of contemporary misinformation environments^13^. The expressed desire for behavioral communication, social media literacy, and scenario-based training in the present study therefore underscores important gaps that future training programs must address.

Overall, this study contributes to the global evidence base by demonstrating that the challenges observed in Lagos State mirror those identified in higher-income and other LMIC contexts. The convergence of findings across settings indicates that effective infodemic management requires both individual-level competencies and system-level strengthening, consistent with global recommendations. By integrating quantitative and qualitative results, this study provides actionable insights that can inform the design of more targeted, practical, and system-supported training models for Nigeria’s PHC workforce and similar settings worldwide.

## Conclusion

This mixed-methods study provides a comprehensive assessment of the infodemic management capacities of frontline health educators and supervisors across three districts in Lagos State, Nigeria. The findings reveal a workforce that is highly aware of the nature and impact of health misinformation and consistently encounters a wide range of rumours during routine community engagements. Despite this strong knowledge base and positive attitudes toward combating misinformation, significant gaps remain in practical skills, confidence, and integration of misinformation response into everyday public health practice.

The qualitative and quantitative data converge to show that these gaps are shaped not only by individual capacity limitations but also by structural and systemic constraints. Entrenched cultural beliefs, distrust in formal health systems, and the rapid spread of misinformation via digital platforms present challenges that exceed what individual educators can address without adequate tools or institutional support. The absence of formal rumour-tracking/management systems, limited supervisory structures, and insufficient access to communication resources further restrict the effectiveness of misinformation response within the primary health care system.

Importantly, the study found no meaningful differences between trained and untrained participants, underscoring the need for training that is targeted, practical, continuous, and grounded in behavioural communication and digital literacy. Frontline educators and supervisors expressed clear expectations for scenario-based learning, digital-media-focused content, and supportive systems that enable coordinated responses to emerging misinformation trends.

Overall, this study highlights the urgent need for a holistic, system-strengthening approach to infodemic management in Lagos State and similar settings. Strengthening workforce competencies must go hand in hand with building institutional structures such as rumour surveillance platforms, supervisory support mechanisms, and coordinated communication processes that empower educators to apply their knowledge effectively. By addressing both individual and systemic gaps, health systems can equip frontline educators to more effectively counter misinformation, build community trust, and enhance resilience against future infodemics.

### Recommendations for Future Research

Future research should explore several areas to deepen understanding of infodemic management within primary health care systems. First, there is a need for longitudinal studies that examine how misinformation trends evolve over time within communities and how frontline educators adapt their strategies in response. Such work would help identify which competencies are most critical for sustained misinformation resilience.

Second, future research should investigate the effectiveness of targeted training interventions, particularly those focused on behavioural communication, digital literacy, and scenario-based skill building. Experimental or quasi-experimental designs could help determine which training formats produce measurable improvements in knowledge, skills, confidence, and integration into routine practice.

Third, additional studies should assess the implementation and impact of structured rumour-tracking systems within PHC settings. Evaluating how digital or hybrid reporting tools influence response speed, coordination, and community trust would offer valuable insights for health system strengthening.

Fourth, research should explore community perceptions and trust dynamics from the perspective of residents themselves. Understanding why certain sources such as religious leaders, peers, or online influencers are perceived as more credible than health workers could inform more effective engagement strategies.

Finally, future studies should examine organizational and policy-level determinants of infodemic management, including leadership practices, resource allocation, and cross-sector collaboration. Investigating these structural factors would support the development of multi-level models that integrate individual competencies with supportive system environments.

Collectively, such research will contribute to a more robust evidence base for designing effective, context-responsive infodemic management frameworks in Nigeria and comparable settings.

## Limitations

This study has several limitations that should be considered when interpreting the findings. First, the study was conducted in three districts within Lagos State, which may limit the generalizability of the results to other states or regions with different sociocultural contexts, health system structures, or misinformation patterns. Although the districts were selected to reflect diverse PHC settings, the findings may not fully capture variations present in rural or hard-to-reach areas.

Second, the quantitative component relied on self-reported data, which may be subject to social desirability bias. Participants may have overestimated their knowledge or confidence, or underreported challenges, particularly given the professional expectations associated with their roles. Although qualitative findings helped contextualize these responses, self-report bias cannot be fully eliminated.

Third, the cross-sectional design captures insights at a single point in time, limiting the ability to observe changes in misinformation dynamics, workforce competencies, or system responsiveness over time. Given the rapidly evolving nature of infodemics especially on digital platforms the findings may not reflect future trends or emerging rumours.

Fourth, while qualitative data provided rich insights into experiences and perceptions, participation in FGDs and KIIs depended on availability and willingness, which may have influenced the range of perspectives captured. Those who chose to participate might differ from those who did not, potentially introducing selection bias.

Finally, the study did not independently verify the content or prevalence of rumours circulating in communities; instead, it relied on participants’ descriptions. Although these accounts were consistent and credible, future research incorporating social listening or community-level data collection could enrich and validate these findings.

Despite these limitations, the mixed-methods approach strengthened the study by allowing triangulation of findings, enhancing the credibility and depth of the conclusions drawn.

## Appendix A: Health Educators’ Infodemic Management Training Needs Assessment Questionnaire (HEIM-TNAQ)

The HEIM-TNAQ is a 30-item instrument assessing frontline health educators’ competencies across five domains: Knowledge, Decision-Response Skills, Attitudes, Confidence, and Integration into Routine Practice.

### Background Information

`o` Age range
`o` Sex (Gender)
`o` Highest educational qualification
`o` Designation
`o` Years of experience in health education/health promotion

### Section A: Knowledge of Health Misinformation (6 items)

**Response format:** Correct = 1, Incorrect = 0

1. Health misinformation refers to:

A. Accurate health information shared widely
B. False or misleading information related to health
C. Official government health announcements
D. Scientific research publications
2. An *infodemic* is best defined as:

A. A shortage of health information
B. An overabundance of information including misinformation that spreads rapidly
C. A new infectious disease outbreak
D. A vaccination campaign
3. Which of the following channels most commonly contributes to the rapid spread of health misinformation?

A. Scientific journals
B. Official government websites
C. Social media and messaging platforms
D. Medical textbooks
4. The first step when encountering suspicious health information is to:

A. Share it immediately with colleagues
B. Verify the source and credibility of the information
C. Ignore it completely
D. Save it for later use
5. Which of the following is considered the most reliable source for public health information?

A. Anonymous social media posts
B. National public health authorities
C. Personal blogs
D. Unverified community rumours
6. Health rumours during outbreaks can influence:

A. Community health-seeking behaviour
B. Agricultural production
C. Weather conditions
D. Transportation systems

### Section B: Decision-Response Skills (Scenario-Based) (6 items)

**Response format:** Correct = 1, Incorrect = 0

1. During a community meeting, someone claims that vaccines cause infertility because they saw a video online. What should you do first?

A. Ignore the statement
B. Ask questions and provide evidence-based clarification
C. End the discussion immediately
D. Criticize the person publicly
2. A community member says malaria is caused by spiritual attacks. What would be the best response?

A. Agree with the explanation
B. Provide a scientific explanation about malaria transmission
C. Change the topic
D. Ignore the claim
3. A WhatsApp message claims that a herbal mixture cures cholera. What should you do?

A. Encourage people to try the remedy
B. Verify the claim and explain the need for medical treatment
C. Forward the message to colleagues
D. Ignore it
4. A religious leader discourages vaccination in the community. What approach is most appropriate?

A. Publicly confront the leader
B. Engage the leader respectfully and provide accurate information
C. Cancel vaccination activities
D. Ignore the issue
5. A rumour spreads that a disease outbreak is fake. What should you do?

A. Ignore the rumour
B. Provide verified information and engage the community
C. Wait for the rumour to disappear
D. Share the rumour to warn others
6. Community members distrust official health information. What strategy would be most effective?

A. Repeat the same messages repeatedly
B. Engage trusted community leaders and explain the evidence
C. Avoid discussing the topic
D. Stop community outreach

### Section C: Attitudes Toward Addressing Health Misinformation (6 items)

**Response scale:**

1 = Strongly disagree, 2 = Disagree, 3 = Neutral, 4 = Agree, 5 = Strongly agree (Negatively worded items are reverse-coded)

1. Addressing health misinformation should be part of the responsibilities of health educators.
2. Health misinformation can significantly influence community health behaviours.
3. Health educators should actively correct misinformation during community engagement activities.
4. Monitoring rumours circulating in communities is important for effective health communication.
5. Addressing misinformation is *not* an important responsibility of health educators. *(Reverse coded)*
6. Collaboration with community leaders is important when addressing misinformation.

### Section D: Confidence in Responding to Health Misinformation (6 items)

**Response scale:**

1 = Not confident, 2 = Slightly confident, 3 = Moderately confident, 4 = Confident, 5 = Very confident

(Negatively worded items are reverse-coded)

1. I feel confident identifying misinformation circulating in communities.
2. I feel confident correcting misinformation during community meetings.
3. I feel confident explaining complex health information in simple terms.
4. I feel confident engaging individuals who strongly believe misinformation.
5. I find it difficult to respond when community members insist on misinformation. *(Reverse coded)*
6. I feel confident responding to misinformation during disease outbreaks.

### Section E: Integration into Routine Practice (6 items)

**Response scale:**

1 = Never, 2 = Rarely, 3 = Sometimes, 4 = Often, 5 = Always (Negatively worded items are reverse-coded)

1. I document rumours or misinformation encountered during community outreach.
2. I report misinformation trends to supervisors or colleagues.
3. I address misinformation during routine health education sessions.
4. I monitor community discussions to identify emerging rumours.
5. Rumours heard during community outreach are usually not worth documenting. *(Reverse coded)*
6. I verify health information using trusted sources before sharing it with communities.

### Domain Structure and Scoring Summary

| Domain | Items | Scoring |
| --- | --- | --- |
| Knowledge | 1–6 | Correct = 1, Incorrect = 0 |
| Decision-Response Skills | 7–12 | Correct = 1, Incorrect = 0 |
| Attitudes | 13–18 | Likert 1–5 (with reverse coding) |
| Confidence | 19–24 | Likert 1–5 (with reverse coding) |
| Integration | 25–30 | Likert 1–5 (with reverse coding) |

### Appendix B: Focus Group Discussion (FGD) Guide

This guide was used to explore frontline health educators’ experiences with health misinformation, their response practices, and their perceived training and support needs. The FGD format is semi-structured, allowing facilitators to probe for further detail as needed.

### Section 1: Introduction and Warm-Up

**Purpose:**

To welcome participants, explain the session objectives, and build rapport.

**Guide Script:**

- Welcome participants and introduce the facilitator/note taker.
- Explain the purpose of the study: understanding experiences with misinformation and training needs.
- Assure confidentiality and voluntary participation.
- Request consent for audio-recording.
- Invite participants to introduce themselves (name, role, years of experience).

### Section 2: Experiences With Health Misinformation Primary Question 1

**“What types of health misinformation do you commonly hear in the communities where you work?”**

**Probes:**

- Misinformation about vaccines
- Misinformation about disease causes
- Misinformation about treatments or herbal remedies
- Outbreak-related rumours (e.g., disbelief in outbreaks)
- Who typically spreads these rumours?

### Section 3: Sources and Channels of Misinformation Primary Question 2

**“Where do these rumours or misinformation usually come from?” Probes:**

- Digital platforms (WhatsApp, Facebook)
- Community conversations (markets, meetings, gatherings)
- Religious and traditional leaders
- Family and peer networks
- Media sources (radio, print, etc.)

### Section 4: Response Strategies Used by Health Educators Primary Question 3

**“How do you usually respond when community members share misinformation with you?” Probes:**

- Dialogue and Q&A approaches
- Use of simplified explanations
- Correcting misinformation immediately
- Involving community leaders
- Escalation to supervisors
- Situations where response was difficult

### Section 5: Challenges in Addressing Misinformation Primary Question 4

**“What challenges do you face when responding to misinformation in the community?” Probes:**

- Entrenched beliefs
- Distrust in government or health workers
- Rapid spread through social media
- Emotional or defensive reactions
- Limited time, tools, or support
- Challenges involving influential figures

### Section 6: Training Experiences and Gaps

**Primary Question 5:**

“Have you received any training related to misinformation response? If yes, what did it cover?”

**Probes:**

- Type and duration of training
- Relevance to real-world misinformation
- Gaps in existing training
- Desired improvements

### Section 7: Training Needs and Preferences

**Primary Question 6:**

**“What type of training would help you respond better to misinformation?”**

**Probes:**

- Behavioural/communication skills
- Social media literacy
- Scenario-based or simulation training
- Refresher courses
- Preferred training format (in-person, blended, online)

### Section 8: Support Systems and Organisational Needs

**Primary Question 7:**

**“What support systems would help health educators manage misinformation better?” Probes:**

- Rumour-reporting platforms
- Access to updated information
- Peer learning
- Supervisory support
- Communication materials

### Section 9: Evaluation of Health Education Impact

**Primary Question 8:**

**“How is the impact of your health education activities currently evaluated?” Probes:**

- Existing indicators
- Gaps in evaluation
- Misinformation-related indicators
- Challenges in monitoring outcomes

### Section 10: Closing Closing Script

- Thank participants for their time and insights.
- Summarize key discussion points.
- Remind participants of confidentiality.
- Provide contact details for follow-up questions.
- Offer an opportunity for any final comments.

### Appendix C: Key Informant Interview (KII) Guide

This semi-structured KII guide was used to explore the perspectives of supervisors, programme managers, and coordinators overseeing health education and community engagement activities. It focuses on system-level insights related to misinformation, workforce preparedness, and organisational support needs.

### Section 1: Introduction

**Purpose:**

To orient the participant, explain objectives, and build rapport.

### Guide Script

- Introduce yourself and the purpose of the interview.
- Clarify that the discussion focuses on their supervisory or managerial experience with misinformation.
- Explain confidentiality and voluntary participation.
- Request permission to audio-record.
- Confirm the participant’s role, district, and years of supervisory experience.

### Section 2: Types of Misinformation Encountered Primary Question 1

**“From your perspective as a supervisor/manager, what types of health misinformation are commonly circulating in the communities?”**

**Probes:**

- Vaccine-related misinformation (e.g., infertility, population control)
- Misinformation about disease causes
- Treatment-related misinformation (e.g., preference for herbs)
- Outbreak-related denial
- How misinformation reaches the health system’s attention

### Section 3: Sources and Channels of Misinformation Primary Question 2

**“Where do these rumours or misinformation typically originate from?” Probes:**

- Social media (WhatsApp, Facebook, viral voice notes)
- Traditional leaders
- Religious gatherings
- Markets and community meetings
- Print, radio, or other media
- Influence of trusted community figures

### Section 4: How Supervisors Respond to Misinformation Primary Question 3

**“How do you and your team typically respond when misinformation is identified?” Probes:**

- Guidance given to health educators
- Corrective actions or clarifications
- Additional community sensitisation meetings
- Collaboration with community leaders
- Coordination with LGA or state health teams
- Examples of successful or challenging response scenarios

### Section 5: Challenges Faced at Supervisory Level Primary Question 4

**“What challenges do you face as a supervisor when addressing misinformation in the communities?”**

**Probes:**

- Entrenched beliefs
- Distrust in government or official sources
- Rapid spread through digital platforms
- Limited tools or systems for monitoring rumours
- Capacity gaps among health educators
- Resource constraints (staffing, materials, transport)

### Section 6: T**raining Exposure and Gaps**

**Primary Question 5:**

**“Have you or your staff received any training related to misinformation, risk communication, or infodemic management?”**

**Probes:**

- Relevance of previous training
- Content gaps
- Adequacy of duration, frequency, and format
- Whether training improves practice
- How training needs differ between supervisors and frontline educators

### Section 7: Training Needs and Capacity Strengthening Primary Question 6

**“What types of training would better prepare health educators and supervisors to manage misinformation?”**

**Probes:**

- Behavioural communication strategies
- Social-media monitoring and response
- Scenario-based or simulation exercises
- Supportive supervision techniques
- Content updates during outbreaks
- Preferred training modalities

### Section 8: Organisational and System-Level Support Needs Primary Question 7

**“What additional support systems or structures would help your team respond more effectively to misinformation?”**

**Probes:**

- Rumour-reporting and tracking platforms
- Access to communication materials (FAQs, flyers, scripts)
- Supervisory support systems
- Peer-learning forums across districts
- Coordination between PHC, LGA, and state levels
- Integration into routine monitoring systems

### Section 9: Evaluation and Monitoring Primary Question 8

**“How is the impact of your health education or misinformation-response activities currently evaluated?”**

**Probes:**

- Existing monitoring indicators
- Gaps in measuring misinformation reduction
- Need for new evaluation tools
- Use of feedback loops
- Reporting pathways and barriers

### Section 10: Closing Closing Script

- Summarize key themes discussed.
- Ask if the participant has additional comments.
- Thank them sincerely for their participation and valuable insights.
- Provide contact information for follow-up questions.

13. World Health Organization. (2025). Impact of the COVID-19 infodemic on frontline workers and health systems: Analysis of story-telling approach for infodemic management. WHO Press.

## Data Availability

The data underlying the findings of this study are available from the corresponding author upon reasonable request

## References

1. Ishizumi A, Kolis J, Abad N, Prybylski D, Brookmeyer KA, Voegeli C, et al. Beyond misinformation: developing a public health prevention framework for managing information ecosystems. The Lancet Public Health. 2024 Apr 20;9(6):e397–e406.

2. Fridman I, Johnson S, Elston Lafata J. Health Information and Misinformation: A Framework to Guide Research and Practice. JMIR Med Educ. 2023 June 7;9:e38687.

3. Aslan D, Kamberi F, Yegenoglu S. Editorial: Infodemic management in public health crises. Front Public Health. 2024 Dec 17;12.

4. Ma X, Vervoort D, Reddy CL, Park KB, Makasa E. Emergency and essential surgical healthcare services during COVID-19 in low-and middle-income countries: A perspective. International Journal of Surgery. 2020 May 16;79(9):43–46.

5. Kilmarx PH, Maitin T, Adam T, Akuffo H, Aslanyan G, Cheetham M, et al. A Mechanism for Reviewing Investments in Health Research Capacity Strengthening in Low-and Middle-Income Countries. Annals of Global Health. 2020 Aug 3;86(1).

6. Topp SM, Abimbola S, Joshi R, Negin J. How to assess and prepare health systems in low-and middle-income countries for integration of services-a systematic review. Health policy and planning. 2017 Dec 18;33(2):298–312.

7. Malik MA. Vaccines and Misinformation: A Public Health Battle. AIJCTH. 2025 Dec 19;1(4):42–51.

8. Verhagen LM, De Groot R, Lawrence CA, Taljaard J, Cotton MF, Rabie H. COVID-19 response in low-and middle-income countries: Don’t overlook the role of mobile phone communication. International Journal of Infectious Diseases. 2020 Aug 4;99(7):334–337.

9. Hanlon C, Luitel NP, Kathree T, Murhar V, Shrivasta S, Medhin G, et al. Challenges and Opportunities for Implementing Integrated Mental Health Care: A District Level Situation Analysis from Five Low-and Middle-Income Countries. PLoS ONE. 2014 Feb 18;9(2):e88437.

10. Introne J, Mckernan B, Corsbie-Massay CLP, Rohlinger D, Tripodi FB. Healthier information ecosystems: A definition and agenda. Asso for Info Science & Tech. 2024 Aug 8;75(10):1025–1040.

11. Pope J, Byrne P, Devane D, Purnat TD, Dowling M. Health misinformation: protocol for a hybrid concept analysis and development. HRB Open Res. 2022 Nov 1;5:70.

12. Padalko H, Chomko V, Yakovlev S, Chumachenko D. A Novel Comprehensive Framework for Detecting and Understanding Health-Related Misinformation. Information. 2025 Feb 26;16(3):175.

13. Rubinelli S, Purnat TD, Wilhelm E, Traicoff D, Namageyo-Funa A, Thomson A, et al. WHO competency framework for health authorities and institutions to manage infodemics: its development and features. Hum Resour Health. 2022 May 7;20(1).

14. Purnat T, Bertrand-Ferrandis C, Yau B, Ishizumi A, White B, Briand S, et al. Training health professionals in infodemic management to mitigate the harm caused by infodemics. The European Journal of Public Health. 2022 Oct 21;32(Suppl 3).

15. Feroz A, Kadir MM, Saleem S. Health systems readiness for adopting mhealth interventions for addressing non-communicable diseases in low-and middle-income countries: a current debate. Global Health Action. 2018 Jan 1;11(1):1496887.

16. Pallas SW, Minhas D, Pérez-Escamilla R, Taylor L, Curry L, Bradley EH. Community health workers in low-and middle-income countries: what do we know about scaling up and sustainability? Am J Public Health. 2013 May 16;103(7):e74–e82.

17. Abuhaloob L, Purnat TD, Tabche C, Atwan Z, Dubois E, Rawaf S. Management of infodemics in outbreaks or health crises: a systematic review. Front Public Health. 2024 Mar 15;12.

18. Utunen H, Balaciano G, Arabi E, Tokar A, Bhatiasevi A, Noyes J. Learning interventions and training methods in health emergencies: A scoping review. PLoS ONE. 2024 July 16;19(7):e0290208.

19. Kyabaggu R, Marshall D, Ebuwei P, Ikenyei U. Health Literacy, Equity, and Communication in the COVID-19 Era of Misinformation: Emergence of Health Information Professionals in Infodemic Management. JMIR Infodemiology. 2022 Apr 28;2(1):e35014.

